# The lasting impact of NICU admission on the parents of children with non-cardiac congenital anomalies: trauma, mental health, and unmet support needs 5-16 years following discharge

**DOI:** 10.64898/2026.06.24.26354566

**Authors:** Laura J. Wilkie, Stephanie Malarbi, Nicholas P. Ryan, Amanda G. Wood

## Abstract

**Background:** Despite growing evidence that interventions targeting parental distress are associated with improved outcomes in families of children with life-threatening conditions, mental health research is limited for parents of NICU graduates treated for non-cardiac congenital anomalies.

**Aims:** To examine the prevalence and severity of mental health difficulties and post-traumatic stress, including subthreshold trauma-related distress, in these parents.

**Method:** Participants were 103 parents (*n*=86 female) of children, aged 5-16 years, who were treated in the NICU for non-cardiac congenital anomalies (e.g., congenital diaphragmatic hernia [CDH], tracheo-oesophageal fistula and/or oesophageal atresia [TOF-OA], abdominal wall defects) at a large tertiary-level paediatric hospital in Australia. Validated measures of mental health (DASS-21) and post-traumatic stress (PCL-5) were administered using an online cross-sectional survey. Whole group and diagnostic subgroup scores were compared with normative data. Comparisons between parents of primary school and high school-aged children enabled the examination of differences in unmet support needs according to their child’s developmental stage.

**Results:** Seventy-four percent of parents reported experiencing mental health difficulties since their child’s congenital anomaly diagnosis, yet only 44.7% had accessed professional mental health support. The mean DASS-21 ‘Stress’ score was significantly elevated relative to Australian population norms (*p*<0.0005). Scores on the PCL-5 indicated that 9.4% met DSM-5 criteria for provisional PTSD diagnoses and a further 20.8% met subthreshold PTSD criteria. Importantly, 50% of parents reporting subthreshold PTSD had not accessed professional psychological support. Mental health concerns appeared more prominent among parents of children with TOF-OA and CDH, as well as parents of high school-aged children.

**Conclusions:** Parents report elevated stress and clinically meaningful subthreshold PTSD symptoms long after their child’s NICU discharge, yet many do not access formal support. These findings highlight the importance of trauma-informed approaches to ongoing mental health surveillance and support for parents of NICU graduates with non-cardiac congenital anomalies.

## Introduction

Parent mental health plays a fundamental role in influencing child development, with parent/caregiver mental health difficulties being consistently associated with an increased risk of social, emotional, and behavioural challenges in children [1]. This relationship is especially pronounced in high-stress contexts, including the neonatal intensive care unit (NICU), where parents of critically ill infants often experience elevated levels of anxiety, depression, and acute stress [2]. For some of these parents, mental health challenges persist long after their child’s initial hospitalisation, with potential implications for parent-child attachment and their child’s long-term psychological functioning [3,4]. Interventions targeting parental distress are reported to reduce anxiety and post-traumatic stress symptoms (PTSS) in parents of children with a range of life-threatening illnesses, with associated improvements in their child’s mental health outcomes [5,6]. Despite this, parent-focused mental health support is not routinely integrated into clinical follow-up care for families of NICU graduates.

While the literature has primarily focused on parents of children with congenital heart disease (CHD) and preterm infants [7,8], studies examining mental health in parents of NICU graduates with non-cardiac congenital anomalies remain scarce. Comparisons between existing studies are further limited by substantial methodological heterogeneity, including wide variations in child age at assessment, treatment protocols, comorbidities, and outcome measurement approaches [9]. For example, inconsistent findings have been reported for parents of children with tracheo-oesophageal fistula and/or oesophageal atresia (TOF-OA), a congenital anomaly characterised by the abnormal development of the oesophagus and trachea. While some studies have described elevated levels of anxiety, depression, and PTSS in these parents, others have reported mental health outcomes comparable with the general population [10,11,12,13]. Further research is needed to better understand the psychological support needs of these parents across different stages of their child’s development, particularly to inform targeted, family-centred approaches to mental health surveillance and intervention.

Accordingly, the aim of the current study was to examine the prevalence and severity of mental health difficulties and PTSS in parents of NICU graduates treated for non-cardiac congenital anomalies, aged 5-16 years. A secondary aim was to identify gaps between mental health needs and service utilisation in the Australian context by exploring the proportion of parents with elevated symptoms who had not accessed professional psychological support. Additionally, mental health outcomes were compared between parents of primary school-aged children and parents of high school-aged children. This allowed the identification of differences in parent psychological support needs specific to their child’s developmental stage, recognising that children themselves can exhibit different emotional and behavioural difficulties across childhood and adolescence, both within this high-risk group and the general population [14,15]. Based on the existing literature, it was hypothesised that parents of both primary and high school-aged children would exhibit elevated levels of depression, anxiety, stress, and PTSS on validated measures of mental health and post-traumatic stress disorder (PTSD). Given that psychological support is not routinely integrated into the clinical follow-up of these families, it was further hypothesised that a significant proportion of parents with elevated symptoms would not have accessed psychological support.

## Method

### Study design, participants and setting

This cross-sectional study was conducted at the Royal Children’s Hospital (RCH), a large tertiary-level paediatric hospital located in Melbourne, Australia. The RCH provides care to children from Victoria, Tasmania, and southern New South Wales, alongside medically complex cases from other Australian states and overseas. Parents of children enrolled in the hospital’s Neonatal Neurodevelopmental Follow-Up Clinic (NNFC) were invited to participate in an online mental health survey if they met the following study inclusion criteria:

i. Parent or legal guardian (‘parent’ used henceforth) of a child, aged 5-16 years, who was treated in the RCH NICU before six months of age for one of the following non-cardiac congenital anomalies between 2007 and 2019: TOF-OA, gastroschisis, exomphalos, congenital diaphragmatic hernia (CDH), Pierre Robin Sequence (PRS), or Vein of Galen aneurysmal malformations (VGAM)
ii. Aged 18 years or older at the time of study enrolment
iii. Proficient in the English language.

Parents of the same child were both enrolled onto the study provided they were eligible and individually gave consent to participate. Families of deceased children, as well as those with a known history of family violence or current intervention (legal protection) order in place, were not approached for this study. Eligibility was determined using information from the child’s medical record and the NNFC database.

### Recruitment and data collection

Letters of invitation were mailed to all eligible parents using contact details obtained from their child’s medical record. Parents interested in taking part in the study were invited to access an online participant information and consent form using the weblink or QR code provided on the letter. Once written informed consent was obtained electronically, participants were automatically redirected to the online mental health survey. All study data were collected and managed using REDCap electronic data capture tools hosted at Murdoch Children’s Research Institute [16,17]. Follow-up recruitment phone calls were conducted with non-respondents after a period of two weeks. Figure 1 shows the flow of recruitment and outcome of follow-up calls.

**Figure 1.**
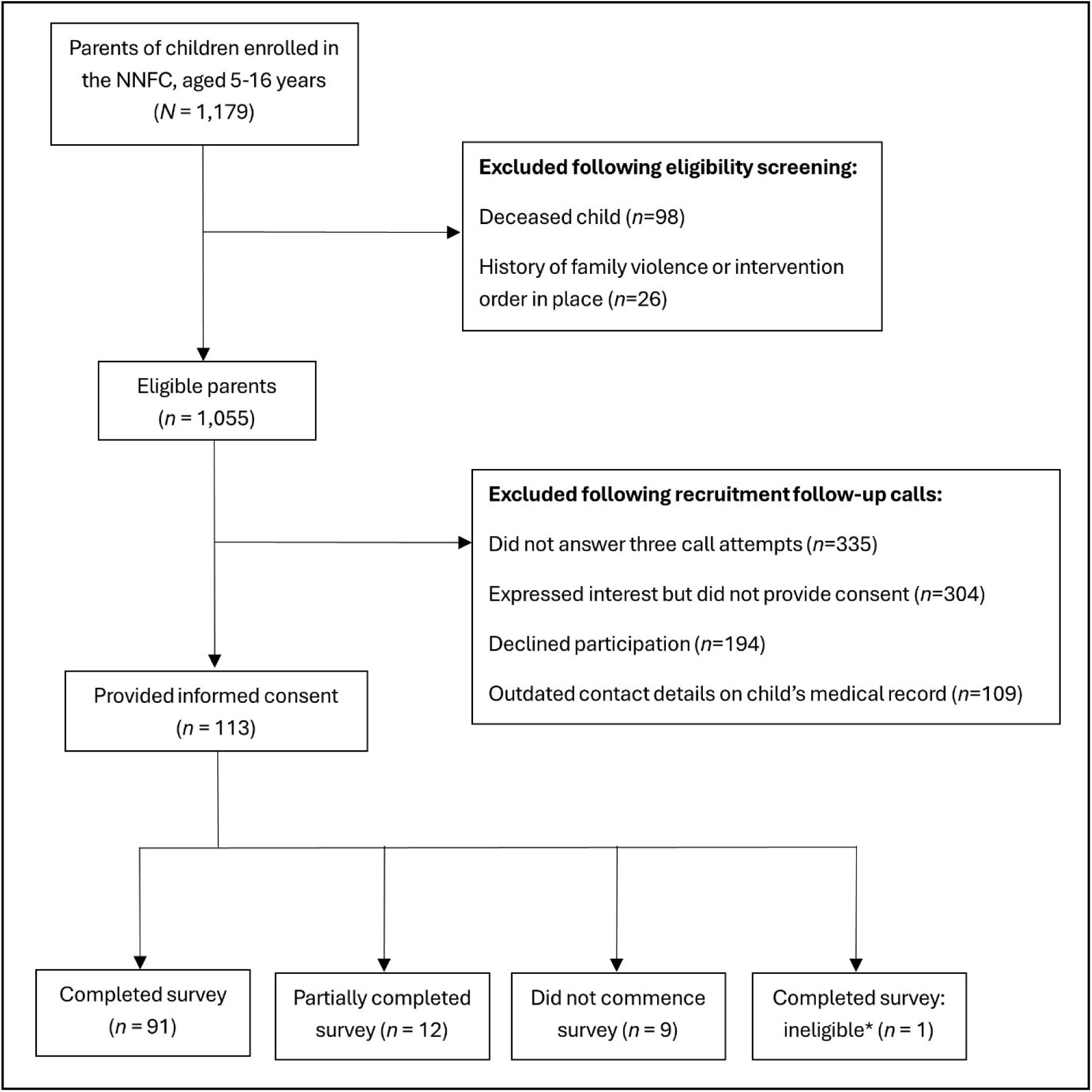
Participant flow diagram. * Participant reported experiencing an adverse life event on the LEC in the two months prior to study enrolment.

### Survey Measures

#### Background information

Clinical and sociodemographic information corresponding to both the participant and their child were obtained from survey responses and the child’s medical record. Background information was selected based on findings from our recent systematic review [9], clinical advice from a consultant neonatologist (LH), and available data. Parents’ sociodemographic data included age, sex, current education level (dichotomised as ‘high school level or below’ and ‘beyond high school level’), country of birth (dichotomised as ‘Australia’ and ‘other’), and current occupational status (categorised as ‘employed’, ‘unemployed’, and ‘retired’). The following retrospective mental health information was also collected from participants: mental health difficulties experienced since their child’s congenital anomaly diagnosis, formal mental health diagnoses, mental health support sought/received since their child’s diagnosis, and the timing of mental health support sought/received.

Clinical and sociodemographic data collected for participants’ children included age, sex assigned at birth, congenital anomaly diagnosis, gestational age at birth (weeks), preterm birth (<37 weeks), birthweight (g), small for gestational age (SGA; defined as birthweight <10^th^ percentile for gestational age and sex according to Australian birthweight centiles [18]), length of first admission (days), days to first medical procedure, cranial ultrasound abnormalities during NICU admission (yes/no), tube fed at NICU discharge (yes/no), comorbid congenital anomalies (yes/no), parent-reported neurodevelopmental diagnoses (yes/no; e.g., autism, attention-deficit/hyperactivity disorder, specific learning disorder), genetic diagnoses (yes/no), current number of caregivers in the home, current school level (dichotomised as ‘primary school’ and ‘high school’), and Index of Relative Socio-economic Advantage and Disadvantage (IRSAD) decile at birth (a proxy of socioeconomic status based on 2011 Australian Census data for postcode [19]).

### Parent mental health

The Depression Anxiety Stress Scales (DASS-21; [20]) consist of three scales containing seven items each (Depression, Anxiety, and Stress). Items are rated on a 4-point Likert scale (0 = ‘never’, 3 = ‘almost always’). A Total Distress score is calculated by summing all DASS-21 responses and multiplying by two. Higher scores indicate more severe symptoms. Cronbach’s α = .88 depression, .82 anxiety, and .90 stress [21]. Scores on the DASS-21 for Depression, Anxiety, Stress, and Total Distress were compared to Australian population norms [22] and categorised by severity level (‘normal’, ‘mild’, ‘moderate’, ‘severe’, and ‘extremely severe’) according to established cut-offs [20]. Scores in the ‘mild’ range or above were considered ‘elevated’.

### Parent post-traumatic stress

The PTSD Checklist for DSM-5 with Life Events Checklist (LEC) and Criterion A (PCL-5; [23]) contains 20 items with four domains corresponding to the four PTSD symptom clusters outlined in the DSM-5: re-experiencing (Criterion B), avoidance (Criterion C), negative alterations in cognition and mood (Criterion D), and hyperarousal (Criterion E). The type and severity of PTSS are determined through the rating of items on a 5-point Likert scale (0 = ‘not at all’, 4 = ‘extremely’), with higher scores indicating more severe PTSS. Cronbach’s α = .94 total symptom severity score [24]. As per the PCL-5 manual, participants were considered to meet DSM-5 criteria for provisional PTSD diagnosis if they endorsed the following criteria in the ‘moderate’ range or above: at least one Criterion B item, one Criterion C item, two Criterion D items, and two Criterion E items. Based on previous recommendations [25], participants who met two or three of the DSM-5 criteria were considered to report symptoms consistent with subthreshold PTSD. Participants were asked to consider their child’s condition as the traumatic experience when completing the PCL-5. The additional completion of the LEC and Criterion A helped determine whether participants had been exposed to other stressful events throughout their life that may impact the outcome of the PCL-5 measure.

### Statistical Analyses

Data were analysed using the IBM Statistical Package for the Social Sciences (SPSS) Version 30.0 (IBM Corp., Armonk, NY, USA). Bonferroni correction was applied to adjust the alpha value for multiple comparisons. Continuous data were presented as means (M) and standard deviations (SD) for normally-distributed data, and medians and interquartile ranges (IQR) for non-normal data, unless specified otherwise. Categorical data were presented as frequencies and percentages. Independent samples *t*-tests, Mann-Whitney U tests, Pearson Chi-Square tests and Fishers Exact tests were computed as appropriate to examine group differences in clinical and sociodemographic characteristics between parents of primary school and high school-aged children.

Given prior evidence of mental health challenges in parents of NICU graduates, hypotheses were directional, supporting the use of one-tailed tests for comparisons with normative data. One-sample *t*-tests were computed to compare mean DASS-21 scores to Australian population norms for the whole group and diagnostic subgroups (excluding VGAM due to extremely small sample size). Effect sizes were determined using Cohen’s D and interpreted according to Cohen’s criteria (0.2 = small, 0.5 = medium, 0.8 = large). Mean scores on the DASS-21 were compared between parents of primary school and high school-aged children using ANCOVA to control for significant differences between the groups for sample characteristics.

Scores on the DASS-21 and PCL-5 were categorised according to established severity levels [20,24]. Binomial tests were computed to compare the distribution of ‘normal’ and ‘elevated’ scores on the DASS-21 to corresponding test norms. The distribution of scores on the DASS-21 and PCL-5 were also compared between parents of primary school and high school-aged children using binary logistic regression analyses to control for significant sociodemographic differences between the groups.

Participants who reported experiencing an adverse life event on the LEC within the two months prior to study enrolment were subsequently excluded from all analyses to reduce the risk of misclassifying transient distress as longer-term psychopathology. This approach is consistent with DSM-5 PTSD criteria requiring symptoms to persist for at least one month. Moreover, to account for lifetime exposure to adverse life events, a Mann-Whitney U test was computed to compare the number of stressful events reported on the LEC between (i) parents who met DSM-5 or subthreshold PTSD criteria and (ii) parents who did not meet criteria.

### Ethical considerations

The authors assert that all procedures contributing to this work comply with the ethical standards of the relevant national and institutional committees on human experimentation and with the Helsinki Declaration of 1975, as revised in 2013. All procedures involving human subjects/patients were approved by the RCH Human Research Ethics Committee (103350) and registered by Deakin University Human Research Ethics Committee (2024-222).

## Results

### Sample characteristics

A total of 113 parents provided informed consent to participate in this study. Of these, 91 completed the survey (80.5%), 12 partially completed the survey (10.6%), nine did not commence the survey (7.9%), and one participant reported an adverse life event in the two months prior to study enrolment and was subsequently excluded from the analyses (0.9%). Figure 1 shows a participant flow diagram, with reasons for exclusion at the screening and recruitment stages.

Sample characteristics for the 103 participants (*n=*86 female) who completed at least one survey measure and were included in the analyses are summarised in Table 1. Clinical and sociodemographic information for participants’ children, aged 5-16 years, are also shown (*n*=99 due to the inclusion of four mother-father dyads). Parent age was the only background variable that differed significantly between parents of primary school-aged children (*n*=50) and parents of high school-aged children (*n*=53) after Bonferroni correction. All subsequent comparisons between these groups were thus controlled for parent age.

**Table 1.** Sample and child characteristics: *n* (%)

| Sample Characteristics | Whole Group | Primary School | High School | Primary vs High School |
| --- | --- | --- | --- | --- |
|  | *n* = 103 | *n* = 50 | *n* = 53 | *p* value |
| Age (years) <sup>a</sup> | 44.3 (6.4) | 41.7 (4.8) | 46.8 (6.7) | <0.0005* |
| Female sex | 86 (83.5) | 42 (84.0) | 44 (83.0) | 0.553 |
| Education beyond high school level | 89 (86.4) | 43 (86.0) | 46 (86.8) | 0.567 |
| Country of birth: |  |  |  |  |
| Australia | 78 (75.7) | 31 (62.0) | 47 (88.7) | 0.002<sup>▲</sup> |
| Other | 25 (24.3) | 19 (38.0) | 6 (11.3) |  |
| Occupational status: |  |  |  |  |
| Employed | 97 (94.2) | 47 (94.0) | 50 (94.3) | 0.833 |
| Unemployed | 5 (4.8) | 3 (6.0) | 2 (1.9) |  |
| Retired | 1 (0.9) | 0 | 1 (0.9) |  |
| **Child Characteristics** | ***n* = 99** | ***n* = 48** | ***n* = 51** |  |
| Congenital anomaly diagnosis: |  |  |  |  |
| TOF-OA | 33 (33.3) | 14 (29.2) | 19 (37.3) | 0.280 <sup>d</sup> |
| CDH | 22 (22.2) | 10 (20.8) | 12 (23.5) |  |
| PRS | 18 (18.2) | 13 (27.1) | 5 (9.8) |  |
| Gastroschisis | 15 (15.2) | 6 (12.5) | 9 (17.6) |  |
| Exomphalos | 9 (9.1) | 4 (8.3) | 5 (9.8) |  |
| VGAM | 2 (2.0) | 1 (2.1) | 1 (2.0) |  |
| Age (years) <sup>b</sup> | 12 (5-16) | 9 [7, 10] | 14 [13, 15] | - |
| Female sex assigned at birth | 48 (48.5) | 23 (47.9) | 25 (49.0) | 0.536 |
| Two caregivers in home | 88 (88.9) | 45 (93.8) | 43 (84.3) | 0.120 |
| IRSAD decile at birth <sup>c</sup> | 8 [6, 9] | 8 [6, 9] | 8 [5, 9] | 0.671 |
| Gestational age at birth (weeks) <sup>c</sup> | 38 [36, 39] | 38 [36, 38] | 38 [36, 39] | 0.389 |
| Preterm (<37 weeks) | 27 (27.3) | 13 (27.1) | 14 (27.5) | 0.574 |
| Birthweight (g) <sup>a</sup> | 2903 (802) | 2893 (811) | 2914 (801) | 0.446 |
| SGA | 14 (14.1) | 7 (14.6) | 7 (13.7) | 0.565 |
| Length of stay: first admission (days) <sup>c</sup> | 32 [19, 53] | 41 [22, 62] | 29 [19, 42] | 0.078 |
| Days to first medical procedure <sup>c</sup> | 2 [1, 7] | 5 [1, 31] | 2 [1, 5] | 0.028<sup>▲</sup> |
| Tube fed at NICU discharge | 55 (55.6) | 30 (62.5) | 25 (49.0) | 0.126 |
| Genetic diagnosis | 12 (12.1) | 10 (20.8) | 2 (3.9) | 0.010<sup>▲</sup> |
| Neurodevelopmental diagnosis | 21 (21.2) | 11 (22.9) | 10 (19.6) | 0.437 |
| Cranial ultrasound abnormalities | 18 (18.2) | 9 (18.8) | 9 (17.0) | 0.546 |
| Comorbid congenital anomalies | 38 (38.4) | 25 (52.1) | 13 (25.5) | 0.006<sup>▲</sup> |

<sup>a</sup> M (*SD*)<sup>b</sup> Median (range)<sup>c</sup> Median [IQR]<sup>d</sup> VGAM group excluded\* Bonferroni corrected $\alpha$ value: 0.0005<sup>▲</sup> Unadjusted $\alpha$ value: 0.05
Notes: IRSAD = Index of Relative Socio-Economic Advantage and Disadvantage

### Parent mental health (retrospective)

Participant mental health information is presented in Table 2. Nearly three quarters of the sample (73.8%) reported that they experienced mental health difficulties since their child’s congenital anomaly diagnosis. Parents most frequently reported mental health difficulties within the first year after their child’s NICU discharge (57.3%). Chi-square tests indicated no significant differences in the frequency of reported mental health difficulties between parents of primary and high school-aged children at any stage of their child’s development up to and including primary school age (i.e., before birth, during NICU admission, within the first year after NICU discharge, preschool, and primary school).

**Table 2.** Sample mental health information: *n* (%)

| **Sample mental health information** | **Whole**<br>**Group**<br>***n* = 103** | **Primary**<br>**School**<br>***n* = 50** | **High**<br>**School**<br>***n* = 53** |
| --- | --- | --- | --- |
| Experienced mental health difficulties since child's diagnosis | 76 (73.8) | 36 (72.0) | 40 (75.5) |
| When mental health difficulties were experienced: |  |  |  |
| Before birth (if child diagnosed antenatally) | 32 (31.1) | 14 (28.0) | 18 (33.9) |
| During NICU admission | 43 (41.7) | 20 (40.0) | 23 (43.4) |
| Within the first year after NICU discharge | 59 (57.3) | 28 (56.0) | 31 (58.5) |
| Preschool | 36 (34.9) | 21 (42.0) | 15 (28.3) |
| Primary school | 38 (36.9) | 18 (36.0) | 20 (37.7) |
| High school | 11/53 (20.8) | - | 11 (20.8) |
| Sought/received mental health support since child's diagnosis | 46 (44.7) | 21 (42.0) | 25 (47.2) |
| Type of mental health support sought/received: |  |  |  |
| Psychologist | 29 (28.2) | 16 (32.0) | 13 (24.5) |
| Counselling | 18 (17.5) | 6 (12.0) | 12 (22.6) |
| Medication | 15 (14.6) | 5 (10.0) | 10 (18.9) |
| When mental health support was sought/received: |  |  |  |
| Before birth (if child diagnosed antenatally) | 7 (6.7) | 3 (6.0) | 4 (7.5) |
| During NICU admission | 4 (3.8) | 2 (4.0) | 2 (3.8) |
| Within the first year after NICU discharge | 17 (16.5) | 9 (18.0) | 8 (15.1) |
| Preschool | 20 (19.4) | 13 (26.0) | 7 (13.2) |
| Primary school | 23 (22.3) | 11 (22.0) | 12 (22.6) |
| High school | 8/53 (15.1) | - | 8 (15.1) |
| Formal mental health diagnosis: | 37 (35.9) | 14 (28.0) | 23 (43.4) |
| Anxiety Disorder | 23 (22.3) | 9 (18.0) | 14 (26.4) |
| Depression | 22 (21.4) | 12 (24.0) | 10 (18.9) |
| Post-Traumatic Stress Disorder | 6 (5.8) | 2 (4.0) | 4 (7.5) |

Since their child’s diagnosis, 44.7% of parents had sought or received mental health support, most commonly through psychologist appointments. Mental health support was most frequently accessed when their child was enrolled in primary school. McNemar tests indicated that, at all stages except high school, the number of parents reporting mental health difficulties significantly exceeded those who sought or received support (*p*<0.0005). The greatest disparity between mental health needs and service utilisation occurred in the first year after NICU discharge. Chi-square tests revealed no significant differences between parents of primary and high school-aged children in the number who accessed mental health support at any stage of their child’s development, up to and including primary school age.

Thirty-six percent of participants reported at least one formal mental health diagnosis, with anxiety disorders and depression being the most prevalent conditions. Parents of high school-aged children were 3.35 times more likely to report a formal mental health diagnosis than those of primary school-aged children, although this association did not remain significant following Bonferroni correction (Supplementary Table 1).

### Parent mental health (prospective)

Mean DASS-21 scale scores and the distribution of DASS-21 scores by severity level are presented in Table 3 and 4, respectively. Parents in our sample had a significantly higher mean score on the Stress scale compared to Australian population norms, although the effect size was small, suggesting a modest elevation at the group level. Notably, 31.1% of parents scored within the ‘elevated’ range for Stress. These findings indicate that, while stress was not uniformly high across the sample, approximately one third of parents endorsed sufficiently heightened stress to potentially warrant targeted support. A substantial proportion of parents who reported ‘elevated’ symptoms on the DASS-21 scales had not engaged with formal mental health care. Thirty-percent of participants with ‘elevated’ Anxiety scores had not sought or received any psychological support and 30% had no formal mental health diagnosis. Similarly, among those with ‘elevated’ Depression scores, 29.6% had not accessed support and 37% had no mental health diagnosis. For parents reporting ‘elevated’ Stress, 31.3% had not sought or received mental health support and 40.6% did not have a formal mental health diagnosis.

**Table 3.** DASS-21 mean scores.

| Group | DASS-21 Scale | Sample Data |  | Sample Data vs Normative Data <sup>a</sup> |  |  |  |
| --- | --- | --- | --- | --- | --- | --- | --- |
|  |  | *n* | *M* (*SD*) | *t* | *df* | Effect size (*d*) | *p* value |
| Whole Group | Depression | 103 | 3.09 (3.43) | 1.53 | 102 | 0.15 | 0.064 |
|  | Anxiety |  | 2.12 (2.63) | 1.45 |  | 0.14 | 0.075 |
|  | Stress |  | 5.82 (3.93) | 4.71 |  | 0.46 | <0.0005* |
|  | Total Distress |  | 11.02 (9.05) | 3.05 |  | 0.30 | 0.001<sup>▲</sup> |
| TOF-OA | Depression | 34 | 3.15 (3.39) | 0.99 | 33 | 0.17 | 0.164 |
|  | Anxiety |  | 2.03 (2.98) | 0.57 |  | 0.10 | 0.287 |
|  | Stress |  | 5.94 (3.99) | 2.85 |  | 0.49 | 0.004<sup>▲</sup> |
|  | Total Distress |  | 11.12 (9.42) | 1.74 |  | 0.30 | 0.045<sup>▲</sup> |
| CDH | Depression | 25 | 3.72 (4.01) | 1.44 | 24 | 0.29 | 0.082 |
|  | Anxiety |  | 2.28 (2.49) | 1.08 |  | 0.22 | 0.145 |
|  | Stress |  | 6.16 (4.08) | 2.66 |  | 0.53 | 0.007<sup>▲</sup> |
|  | Total Distress |  | 12.16 (9.58) | 2.01 |  | 0.40 | 0.028<sup>▲</sup> |
| PRS | Depression | 18 | 2.83 (3.75) | 0.29 | 17 | 0.07 | 0.385 |
|  | Anxiety |  | 1.89 (2.52) | 0.25 |  | 0.06 | 0.402 |
|  | Stress |  | 5.56 (4.37) | 1.52 |  | 0.36 | 0.073 |
|  | Total Distress |  | 10.28 (9.74) | 0.86 |  | 0.20 | 0.200 |
| Gastroschisis | Depression | 15 | 3.13 (2.56) | 0.85 | 14 | 0.22 | 0.204 |
|  | Anxiety |  | 2.73 (2.63) | 1.46 |  | 0.38 | 0.083 |
|  | Stress |  | 6.07 (3.86) | 2.08 |  | 0.54 | 0.028<sup>▲</sup> |
|  | Total Distress |  | 11.93 (8.22) | 1.71 |  | 0.44 | 0.054 |
| Exomphalos | Depression | 9 | 1.56 (2.60) | -1.17 | 8 | 0.39 | 0.138 |
|  | Anxiety |  | 0.89 (1.36) | -1.87 |  | 0.62 | 0.049<sup>▲</sup> |
|  | Stress |  | 4.11 (3.02) | 0.12 |  | 0.04 | 0.454 |
|  | Total Distress |  | 6.56 (6.31) | -0.83 |  | 0.28 | 0.215 |
| Primary School | Depression | 50 | 2.76 (3.57) | 0.38 | 49 | 0.05 | 0.354 |
|  | Anxiety |  | 1.92 (2.39) | 0.53 |  | 0.08 | 0.298 |
|  | Stress |  | 5.74 (4.27) | 2.89 |  | 0.41 | 0.003<sup>▲</sup> |
|  | Total Distress |  | 10.42 (9.33) | 1.61 |  | 0.23 | 0.057 |
| High School | Depression | 53 | 3.39 (3.30) | 1.82 | 52 | 0.25 | 0.037<sup>▲</sup> |
|  | Anxiety |  | 2.30 (2.85) | 1.43 |  | 0.20 | 0.079 |
|  | Stress |  | 5.89 (3.62) | 3.82 |  | 0.52 | <0.0005* |
|  | Total Distress |  | 11.58 (8.85) | 2.70 |  | 0.37 | 0.005<sup>▲</sup> |

<sup>a</sup> Australian population norms (*n* = 497 [22])
Depression: 2.57 (3.86)
Anxiety: 1.74 (2.78)
Stress: 3.99 (4.24)
Total Distress: 8.30 (9.83)
\* Bonferroni corrected $\alpha$ value: 0.0005<sup>▲</sup> Unadjusted $\alpha$ value: 0.05

Compared to Australian population norms, parents of primary school-aged children had a higher mean score for Stress, with 32% scoring within the ‘elevated’ range. However, this difference was non-significant after Bonferroni correction. Among parents of high school-aged children, mean Depression, Stress, and Total Distress scores were higher than population norms, yet the only mean score that remained significantly elevated after correction was Stress, with a medium effect size (Table 3). This group also had a higher proportion of ‘elevated’ Anxiety and Total Distress scores compared to parents of primary school-aged children (Supplementary Table 1). While these results suggest that parents of high school-aged children may face greater mental health challenges, between-group differences on the DASS-21 were not significant after adjusting for multiple comparisons.

For all diagnostic subgroups, mean DASS-21 scores and severity distributions were consistent with Australian population norms following Bonferroni correction (Table 3 and 4). Despite this, descriptive patterns suggested higher-than-expected mean Stress scores for parents of children with TOF-OA, CDH, and gastroschisis. Mean Total Distress scores were also elevated for parents of children with TOF-OA and CDH. In contrast, parents of children with exomphalos had a lower (i.e., favourable) mean Anxiety score compared to population norms, although this subgroup was very small and findings should be interpreted cautiously. Overall, these results suggest that parents of children with TOF-OA and CDH may warrant closer monitoring and consideration of targeted psychological support, although more research with larger samples is needed to further clarify diagnosis-specific support needs.

**Table 4.** Distribution of DASS-21 scores: *n* (%)

| Group | *n* | DASS-21 Index | Normal | Mild | Moderate | Severe | Extremely Severe | Test Norms vs Sample Data <sup>a</sup> |
| --- | --- | --- | --- | --- | --- | --- | --- | --- |
|  |  |  |  |  |  |  |  | *p* value |
| Whole Group | 103 | Depression | 76 (73.8) | 10 (9.7) | 13 (12.6) | 1 (1.0) | 3 (2.9) | 0.119 |
|  |  | Anxiety | 83 (80.6) | 4 (3.9) | 12 (11.7) | 2 (1.9) | 2 (1.9) | 0.392 |
|  |  | Stress | 71 (68.9) | 17 (16.5) | 8 (7.8) | 7 (6.8) | 0 | 0.008<sup>▲</sup> |
| TOF-OA | 34 | Depression | 25 (73.5) | 4 (11.8) | 4 (11.8) | 0 | 1 (2.9) | 0.283 |
|  |  | Anxiety | 28 (82.3) | 2 (5.9) | 3 (8.8) | 0 | 1 (2.9) | 0.394 |
|  |  | Stress | 23 (67.6) | 6 (17.6) | 3 (8.8) | 2 (5.9) | 0 | 0.079 |
| CDH | 25 | Depression | 18 (72.0) | 1 (4.0) | 4 (16.0) | 1 (4.0) | 1 (4.0) | 0.260 |
|  |  | Anxiety | 19 (76.0) | 1 (4.0) | 4 (16.0) | 0 | 1 (4.0) | 0.432 |
|  |  | Stress | 18 (72.0) | 2 (8.0) | 2 (8.0) | 3 (12.0) | 0 | 0.260 |
| PRS | 18 | Depression | 13 (72.2) | 3 (16.6) | 1 (5.5) | 0 | 1 (5.5) | 0.322 |
|  |  | Anxiety | 16 (88.9) | 0 | 1 (5.5) | 1 (5.5) | 0 | 0.238 |
|  |  | Stress | 12 (66.7) | 3 (16.6) | 2 (11.1) | 1 (5.5) | 0 | 0.159 |
| Gastroschisis | 15 | Depression | 11 (73.3) | 1 (6.7) | 3 (20.0) | 0 | 0 | 0.390 |
|  |  | Anxiety | 11 (73.3) | 0 | 3 (20.0) | 1 (6.7) | 0 | 0.390 |
|  |  | Stress | 10 (66.6) | 3 (20.0) | 1 (6.7) | 1 (6.7) | 0 | 0.191 |
| Exomphalos | 9 | Depression | 8 (88.9) | 0 | 1 (11.1) | 0 | 0 | 0.407 |
|  |  | Anxiety | 8 (88.9) | 1 (11.1) | 0 | 0 | 0 | 0.407 |
|  |  | Stress | 7 (77.8) | 2 (22.2) | 0 | 0 | 0 | 0.593 |
| Primary School | 50 | Depression | 38 (76.0) | 4 (8.0) | 6 (12.0) | 0 | 2 (4.0) | 0.364 |
|  |  | Anxiety | 41 (82.0) | 2 (4.0) | 6 (12.0) | 0 | 1 (2.0) | 0.364 |
|  |  | Stress | 34 (68.0) | 8 (16.0) | 4 (8.0) | 4 (8.0) | 0 | 0.041<sup>▲</sup> |
| High School | 53 | Depression | 38 (71.7) | 6 (11.3) | 7 (13.2) | 1 (1.9) | 1 (1.9) | 0.128 |
|  |  | Anxiety | 42 (79.2) | 2 (3.8) | 6 (11.3) | 2 (3.8) | 1 (1.9) | 0.500 |
|  |  | Stress | 38 (71.7) | 8 (15.1) | 4 (7.5) | 3 (5.7) | 0 | 0.128 |

<sup>a</sup> Normal vs Elevated (DASS-21 test norms: 79% Normal, 21% Elevated)<sup>▲</sup> Unadjusted $\alpha$ value: 0.05

### Parent post-traumatic stress (prospective)

A summary of PCL-5 scores is presented in Table 5. Nine parents (9.4%) met DSM-5 criteria for provisional PTSD diagnoses. Of these, one had not accessed mental health support (11.1%) and three (33.3%) had no formal mental health diagnosis. An additional twenty parents (20.8%) met criteria for subthreshold PTSD; half of this group (50%) had neither accessed psychological support nor received a mental health diagnosis. The number of lifetime stressful events reported on the LEC did not differ significantly between (i) parents who met DSM-5 or subthreshold PTSD criteria and (ii) parents who did not meet criteria.

**Table 5.** Frequency of PCL-5 scores that met DSM-5 diagnostic criteria: *n* (%)

| Group | *n* | Met DSM-5 criteria |  |  |  | Provisional PTSD diagnosis <sup>a</sup> | Met subthreshold PTSD criteria <sup>b</sup> |
| --- | --- | --- | --- | --- | --- | --- | --- |
|  |  | Criterion B | Criterion C | Criterion D | Criterion E |  |  |
| Whole Group | 96 | 29 (30.2) | 27 (28.1) | 23 (24.0) | 23 (24.0) | 9 (9.4) | 20 (20.8) |
| TOF-OA | 33 | 8 (24.2) | 7 (21.2) | 8 (24.2) | 10 (30.3) | 2 (6.1) | 8 (24.2) |
| CDH | 24 | 10 (41.7) | 9 (37.5) | 8 (33.3) | 8 (33.3) | 3 (12.5) | 6 (25.0) |
| PRS | 16 | 3 (18.8) | 3 (18.8) | 2 (12.5) | 1 (6.3) | 1 (6.3) | 2 (12.5) |
| Gastroschisis | 12 | 6 (50.0) | 6 (50.0) | 3 (25.0) | 2 (16.7) | 1 (8.3) | 4 (33.3) |
| Exomphalos | 9 | 1 (11.1) | 1 (11.1) | 1 (11.1) | 1 (11.1) | 1 (11.1) | 0 |
| Primary School | 49 | 11 (22.4) | 8 (16.3) | 9 (18.4) | 10 (20.4) | 4 (8.2) | 6 (12.2) |
| High School | 47 | 18 (38.3) | 19 (40.4) | 14 (29.8) | 13 (27.7) | 5 (10.6) | 14 (29.8) |

<sup>a</sup> DSM-5 criteria for provisional PTSD diagnosis: scores in the ‘moderate’ range or higher for at least one Criterion B item, one Criterion C item, two Criterion D items, and two Criterion E items.
<sup>b</sup> Subthreshold PTSD criteria: met DSM-5 criteria for two or three criteria [25].

Re-experiencing was the most frequently endorsed PTSD symptom cluster, with 30.2% of parents rating at least one Criterion B item in the ‘moderate’ range or above. Participants meeting subthreshold or DSM-5 PTSD criteria were predominantly parents of children with TOF-OA and CDH, again suggesting that these groups may warrant closer monitoring or further investigation regarding psychological support needs. Moreover, parents of high school-aged children were 6.94 times more likely to report symptoms of avoidance (Criterion C) than parents of primary school-aged children (Supplementary Table 1), although wide confidence intervals and the loss of significance after Bonferroni correction limit interpretations of this finding.

## Discussion

### Whole group findings

Consistent with the limited existing literature [11,26,27], our findings suggest that parents of NICU graduates with non-cardiac congenital anomalies may be at increased risk of long-term mental health challenges, though this risk does not appear to be universal across all diagnostic subgroups. Three quarters of our sample reported experiencing mental health difficulties since their child’s diagnosis, and over one third disclosed a formal mental health diagnosis. The prevalence of reported psychological difficulties in this group was notably high relative to Australian general population estimates, whereby approximately 22% experience mental illness each year [28]. Despite this, a large proportion of parents reporting current symptoms of depression, anxiety, stress, or PTSS had not accessed psychological support, highlighting a clear gap between parent mental health needs and service utilisation. This gap may reflect a combination of practical and psychological barriers, which can include limited service availability, parental time constraints, and caregiver burden, the prioritisation of their child’s care, and potential uncertainty about what constitutes ‘typical’ adjustment [29]. Some data suggest that inadequate mental health education during NICU admission may limit parent knowledge regarding expected emotional responses and indicators for seeking support after the transition to home and beyond [30]. Early, trauma-informed approaches to parent mental health education may be valuable preventative strategies, supporting parents to better identify and address their own needs alongside those of their infant.

Our findings indicate that the first year after NICU discharge is the period of greatest unmet support needs for parents. This period marks a critical transition from intensive hospital care to managing complex medical needs at home and navigating community-based services. During this time, parents assume increased responsibility for their child’s health, leaving them at elevated risk of mental health struggles [31]. High psychological morbidity has previously been documented among mothers of infants with TOF-OA during this transition period, with 38% reporting clinically elevated trait anxiety on the State-Trait Anxiety Inventory (STAI; [32]). These findings illustrate an urgent need for early mental health screening and accessible support pathways for families of infants with non-cardiac congenital anomalies, especially during the transition from NICU to at-home care, when unmet support needs appear to be particularly pronounced.

Stress was the most frequently endorsed concern on the DASS-21, with approximately one-third of parents reporting elevated stress levels. This high prevalence may potentially reflect the ongoing challenges and uncertainties associated with managing their child’s complex health condition, including repeat hospital admissions and medical procedures, and the emergence of neurodevelopmental delays that become increasingly detectable as their child grows older. Children with non-cardiac congenital anomalies often require extensive at-home care, which can significantly disrupt parents’ professional and social lives [33]. As a result, the demands of caregiving may lead to social isolation. Wallace and colleagues found that parents of children with TOF-OA, aged 0-12 years, who perceived themselves as receiving less support for caring reported more anxiety and depressive symptoms on the Hospital Anxiety Depression Scale [26]. Furthermore, mothers of children with CHD were more likely to report low mental health-related quality of life (QOL) on the Short Form-12 Health Survey if they perceived themselves as having poor social support [34].

While elevated stress levels were commonly reported by parents on the DASS-21 in our study, it is important to note that these scores are not diagnostic. Elevated stress may reflect the cumulative impact of ongoing caregiving demands and, in some cases, an adaptive vigilance response consistent with pediatric medical traumatic stress [35], rather than the presence of a clinical disorder. Accordingly, it is important not to pathologise parental responses, but instead to understand whether families would benefit from, or perceive a need for, additional support. Providing families with access to appropriate mental health resources and social support networks may help to mitigate the impact of caregiving-related pressures, improving long-term QOL and wellbeing for the whole family unit.

Approximately one in ten parents met DSM-5 diagnostic criteria for PTSD, consistent with the 11% lifetime prevalence of PTSD within the Australian general population [28]. Subthreshold PTSD was twice as common as PTSD in our sample, yet half of these parents had not sought or accessed psychological support. Research examining subthreshold PTSD is largely limited to military populations. While its prevalence within the Australian general population is unknown, 13.1% of the US general population are reported to meet subthreshold PTSD criteria [36]. Subthreshold PTSD has been linked to impairments in both mental and physical functioning [37], underscoring the importance of early monitoring and intervention for parents even when full diagnostic criteria are not met. The relatively high frequency of subthreshold PTSD symptoms observed in this group suggests that reliance on diagnostic thresholds alone may underestimate clinically meaningful distress, and that symptom-based surveillance may be particularly important in this population, consistent with recommendations based on Kazak’s Integrative Trajectory Model of Pediatric Medical Traumatic Stress [35].

Among the PTSD symptom clusters, re-experiencing was the most frequently endorsed, with many participants reporting intrusive memories, nightmares, and flashbacks. The ongoing demands of caring for a child with a congenital anomaly may re-expose parents to unavoidable reminders of their child’s health struggles. This may amplify feelings of distress and contribute to parental re-experiencing symptoms, although more research is required to further explore this. To address these challenges, it is essential to provide families with trauma-informed care to help manage symptoms and support overall QOL and emotional wellbeing.

### Subgroup findings

Parents of high school-aged children appeared to have poorer overall mental health outcomes compared to parents of primary school-aged children. This is despite no significant differences between groups across the clinical and sociodemographic background variables examined, with the exception of parent age. Specifically, parents of high school-aged children had higher odds of reporting formal mental health diagnoses, and more frequently endorsed symptoms of anxiety, distress, and avoidance. These results suggest that mental health difficulties may intensify or resurface for some parents as their child grows older. This is consistent with previous findings, whereby a significant increase in intrusive stress was reported on the Impact of Event Scale between 6-months postpartum and 9-year follow-up for parents of children with various congenital anomaly diagnoses, including TOF-OA, CDH, and abdominal wall defects [12].

Multiple factors may contribute to an elevated risk of psychopathology in parents of high school-aged children, including reduced support despite ongoing medical needs, concerns associated with transitions to adult care, and evolving social and academic challenges during adolescence. Moreover, the implementation of Newborn and Family-Centred Care approaches in the NICU, which were not widely used historically, may enhance parental resilience. For example, kangaroo care – a form of skin-to-skin contact whereby parents hold their baby against their bare chest – is reported to significantly reduce the occurrence of PTSD in parents at one month post-discharge [38]. Having left the NICU before the widespread adoption of family-centred practices, parents of high school-aged children may have had less access to structured support systems during admission, potentially limiting early protective influences on their mental health. More research is needed to further examine differences in psychological support needs between these parents, ideally through prospective longitudinal design.

Diagnostic subgroup analyses revealed that parents of children with TOF-OA and CDH may be at heightened risk of mental health challenges compared to parents of children with abdominal wall defects and PRS. This could potentially be attributed to increased medical uncertainty, as these groups often face significant long-term sequelae necessitating extensive treatment, including pulmonary and gastrointestinal problems [39,40]. In contrast, parents of children with exomphalos had lower (i.e., favourable) mean anxiety scores compared to the Australian general population, although this finding should be interpreted cautiously given the small subgroup size and predominance of exomphalos minor (78%), which is known to be associated with fewer neurodevelopmental challenges compared to exomphalos major [41]. Further research with larger samples is needed to examine variations both between and within these diagnostic subgroups.

### Clinical implications

Our findings highlight the importance of integrating routine parent mental health screening into the long-term follow-up care of children with non-cardiac congenital anomalies, with particular attention given to families of children with TOF-OA and CDH. Despite a high prevalence of stress and subthreshold PTSD reported among parents, service uptake was low. This underscores a clear need for more accessible mental health services, potentially by embedding support directly within follow-up clinics. The first year after NICU discharge represents a particularly challenging time for these families, offering a critical window for early psychological support and monitoring. Our results therefore emphasise a need for longitudinal, trauma-informed mental health support beginning during or soon after NICU discharge and continuing through key transition points in their child’s life as care needs evolve, alongside improved mental health education during NICU admission.

### Strengths and limitations

This study benefits from a relatively large sample of parents compared with much of the existing literature in this population, allowing for meaningful insights into the mental health challenges faced by families of children with non-cardiac congenital anomalies. The inclusion of various diagnostic subgroups provides broader insight into parent mental health needs across several non-cardiac congenital anomalies, while recognising that subgroup findings should be interpreted with caution given sample sizes. Although the conservative nature of Bonferroni correction meant that small but clinically relevant effects may have been masked, presenting effect sizes and distributions of scores provides a more nuanced understanding of the data than *p*-values alone, offering a clearer picture of the potential clinical impact.

Several factors may have introduced bias to our study. The sample predominantly consisted of mothers, limiting the generalisability of our findings to fathers. Previous research found that significantly more mothers of children with TOF-OA, aged 4-10 years, reported PTSS in the clinical range of the Perinatal PTSD Questionnaire compared to fathers [27], indicating that maternal and paternal support needs may vary. The sample also included no representation of caregivers other than biological parents, despite all legal caregivers being eligible to participate if inclusion criteria were met. Future research should therefore make a concerted effort to capture the unique perspectives of a broader range of caregivers. Moreover, most of our participants reported education beyond high school level and current employment. Our findings may thus not adequately reflect the experiences of parents with lower levels of education, as well as those facing greater financial stress. In a previous study, lower education levels were identified as a significant risk factor for clinically important state anxiety on the STAI in parents of children with various congenital anomalies [12]. This underscores a need for more research that includes families with diverse educational backgrounds. Our survey was also only available in English, likely limiting participation from linguistically diverse families. Additionally, some families were unreachable during the recruitment process due to outdated contact details on their child’s medical record, potentially contributing to selection bias.

While appropriate from an ethical and safeguarding perspective, this study likely underrepresents families experiencing higher levels of adversity and complexity due to the exclusion of those with a current intervention (legal protection) order or history of family violence. Together with the broader sampling considerations noted above, these factors may have resulted in conservative estimates of parent distress and unmet support needs and should be considered when interpreting the prevalence findings. This highlights the importance of conducting targeted research with affected families to help better understand their experiences and inform the development of appropriate, trauma-informed supports. While these limitations are common in clinical research, addressing them in future studies would strengthen generalisability. To minimise bias, future research should ensure a balanced representation of fathers, offer surveys in multiple languages, and aim to recruit a more diverse sample across educational levels and employment status to better capture the experiences of a broader range of families. The inclusion of a matched control group rather than reliance on test norms would also enable more meaningful comparisons that better account for potential confounders, such as differences in socioeconomic status. This is particularly important considering indicators of socioeconomic disadvantage are established risk factors for gastroschisis [42].

### Recommendations for future research

Our findings suggest that, for some parents, mental health difficulties persist for years beyond their child’s initial NICU stay. Longitudinal research is therefore essential to explore how mental health and PTSS evolve for parents across their child’s development, helping to pinpoint optimal timings for psychological support. Future research should also examine the risk and resilience factors that influence parent mental health outcomes, with a focus on identifying modifiable factors that could be targeted by early intervention. Furthermore, understanding the long-term effects of parent mental health on their child’s own mental health and wellbeing will be critical for informing family-centred approaches to mental health care.

## Conclusion

In summary, parents of NICU graduates treated for non-cardiac congenital anomalies reported elevated levels of stress and subthreshold PTSD symptoms long after their child left the NICU. The first year after discharge emerged as a critical period of heightened distress, and parents of children with TOF-OA and CDH appeared to be disproportionately affected by mental health difficulties, as well as parents of high school-aged children. A large proportion of parents endorsing elevated symptoms had not accessed psychological support. These findings illustrate the importance of early, trauma-informed approaches to ongoing mental health surveillance to assist families in managing the challenges of caring for a child born with a complex congenital condition. Symptom-based monitoring, rather than reliance on diagnostic thresholds alone, may be important when identifying parents who could benefit from support.

## Data Availability

Data not available. The data used for this research includes confidential patient information.

## Acknowledgements

The authors thank Dr Leah Hickey, Dr Trisha Prentice, and Jo Brooks at the Royal Children’s Hospital for providing clinical advice for the purposes of this study. The authors also thank the Royal Children’s Hospital Foundation for funding the Neonatal Neurodevelopmental Follow-Up Clinic and the following staff for their contributions establishing or working in the service: Neonatology team - Professor Rod Hunt, Professor Julia Charlton, Dr Leah Hickey, Dr Margaret Morgan, Dr Ruth Armstrong, Dr Penny Kee; Neuropsychology team - Dr Alice Burnett, Dr Stephanie Malarbi, Dr Esther Hutchinson, Dr Anita Chisolm; Allied Health – Sue Greaves, Lisa Kelly, Dani Centorame, Zoe Strang, Ursula Sevil, Emma Lind, Francyne Samara.

