## Supplementary Table 1 for "The lasting impact of NICU admission on the parents of children with non-cardiac congenital anomalies: trauma, mental health, and unmet support needs 5-16 years following discharge"

**Supplementary Table 1.** Comparisons between parents of primary school-aged children and parents of high school-aged children

| Measure | Comparing group distributions (primary vs high school) | OR [a] | 95% CI | *p* value |
|---|---|---|---|---|
| Mental Health Information | Experienced mental health difficulties since child's diagnosis | 1.61 | [0.59, 4.38] | 0.349 |
| | Sought/received mental health support since child's diagnosis | 1.95 | [0.66, 5.79] | 0.229 |
| | Formal mental health diagnosis | 3.35 | [1.29, 8.71] | 0.013▲ |
| DASS-21 | Depression | 2.34 | [0.85, 6.44] | 0.098 |
| | Anxiety | 2.29 | [0.76, 6.94] | 0.140 |
| | Stress | 1.83 | [0.69, 4.81] | 0.224 |
| PCL-5 | Criterion B | 2.08 | [0.76, 5.70] | 0.153 |
| | Criterion C | 6.94 | [2.18, 22.12] | 0.001▲ |
| | Criterion D | 1.76 | [0.59, 5.16] | 0.307 |
| | Criterion E | 2.06 | [0.87, 6.02] | 0.189 |
| | Subthreshold PTSD | 2.50 | [0.77, 8.16] | 0.128 |
| | Provisional PTSD diagnosis | 2.78 | [0.59, 13.13] | 0.198 |
| | **Comparing group means (primary vs high school)** | **F** | **$\eta^2$** | ***p* value** |
| DASS-21 | Depression | 3.79 | 0.04 | 0.054 |
| | Anxiety | 4.16 | 0.04 | 0.044▲ |
| | Stress | 2.16 | 0.02 | 0.145 |
| | Total Distress | 3.95 | 0.04 | 0.050▲ |

[a] Odds for parents of high school-aged children compared to parents of primary school-aged children

▲ Unadjusted α value: 0.05
